# CAREGIVERS’ PERSPECTIVE: SATISFACTION WITH HEALTHCARE SERVICES AT THE PAEDIATRIC SPECIALIST CLINIC OF THE NATIONAL REFERRAL CENTRE IN MALAYSIA

**DOI:** 10.1101/2021.08.03.21261517

**Authors:** Thinakaran Selvarajah, Eiko Yamamoto, Yu Mon Saw, Tetsuyoshi Kariya, Nobuyuki Hamajima

## Abstract

**Background:** The idea of consumer satisfaction is gaining momentum across all business sectors around the world and a satisfaction survey is utilized as an instrument to recognize deficiencies with various facets of services and offers a valuable opportunity for customers to evaluate their experience with healthcare services. A first research performed at a public hospital’s paediatric clinic, which is also the first hospital to adopt the Public-Private-Partnership (PPP) model under the Ministry of Health Malaysia (MoH), with the aim of discovering prevalence and factors affecting the satisfaction of caregivers at the national referral centre.

**Materials and Methods:** Cross-sectional research using the standard self-administered SERVQUAL questionnaire was conducted amongst caregivers accompanying their child to the clinic. It consists of 16 paired statements to evaluate their expectations and experiences with the clinic services. 459 caregivers were involved with a majority being satisfied with the clinic services.

**Result:** The most significant dimensions were “tangibles”, “assurance” and “outcome”. Caregivers from the Indian community, lower household income, and lower educational background demonstrated a higher level of satisfaction.

**Conclusion:** This article suggests that although most caregivers are very satisfied with the services, greater emphasis must be placed on delivering reliable service in response to MoH’s mission to provide quality and integrated people-centred health services in Malaysia.

## INTRODUCTION

Consumer satisfaction plays an increasingly important role in reforming healthcare quality and delivery in general across the United States of America and Europe (Bleich, Özaltin and Murray, 2009). The Integrated People-Centered Health Services (IPCHS) is a global strategy by the World Health Organization (WHO) that proposes a vision focused on providing people-centred and integrated healthcare services. This is a vision described as: “A future in which all people have access to health services that are provided in a way that responds to their personal preferences, are coordinated around their needs and are safe, effective, timely, efficient and of an acceptable quality, throughout their life course” (World Health Organization, 2015). Quality healthcare, in part, means meeting the needs of patients (Vuori, 1991). And patients as the main stakeholders in a healthcare system, their satisfaction reflects the expectations and general experience of healthcare services provided to them (Mukhtar et al., 2013).

Patient satisfaction serves as an objective indicator of experiences, health outcomes, and trust with the health care system, representing whether the care provided has satisfied the patient’s needs and expectations (Larson et al., 2019). Besides, it is an evaluation of the services provided by healthcare providers, which are influenced by both the level of expectations and the patient’s experience (Zun, Ibrahim and Hamid, 2018). It is also possible to monitor the quality of care that could pave ways toward improving health care delivery (Jenkinson et al., 2002). Research suggests that satisfied patients are more prepared to seek medical guidance, comply with therapies, fulfil appointments, and to refer other patients to a physician (Donabedian, 2013; (Health Services Research Group, 1992). A research carried out in India indicates that surveys of patient satisfaction also act as a way to hold doctors responsible (Sharma A., Kasar and Sharma R., 2014). In addition, the advent of increased competitiveness in the healthcare sector has led to the development of facilities that are committed to meeting the needs of patients. Highly ranked institutions in terms of quality of service have better customer retention, lower expenses for bringing in new customers, increased profitability as well as higher customer satisfaction (Cronin, Brady and Hult, 2000; Janda, Trocchia and Gwinner, 2002; Gounaris and Dimitriadis, 2003; Yoon and Suh, 2004).

The Ministry of Health Malaysia (MoH) began its quality assurance program since the 1980s and has implemented many initiatives to improve the quality of healthcare delivery and enhance customer satisfaction, which includes the Client’s Charter and the acculturation of corporate values among employees who are caring, professional and exercise teamwork (Abd Manaf, 2012). Malaysia has provided impressive health benefits for its population, through low-cost healthcare funded primarily by general revenue and taxes collected by the federal government (World Health Organization. Regional Office for the Western Pacific, 2012). The government has continuously committed itself to healthcare equity and accessibility, with the public health sector financing almost 95% of the cost of treatment and subsequently providing access to healthcare for more than 90% of its population (Ministry of Health Malaysia, 2011). Malaysians are also granted free access to consultations, treatment, and medications, as both inpatients and outpatients, for a nominal registration fee of Malaysian Ringgit (RM) 1.00 (USD 0.33), in all public healthcare facilities in the country (Ganasegeran et al., 2015). This long-standing public policy has instilled a sense of entitlement among Malaysians that healthcare services in Malaysia should be free or cost the very least (World Health Organization. Regional Office for the Western Pacific, 2012).

Many studies conducted at public healthcare facilities in Malaysia have shown a high level of patient satisfaction with the services provided (Kaur et al., 2019). However, to our best knowledge, no studies have been conducted on caregivers’ satisfaction in MoH paediatric outpatient clinics or facilities. This study is, therefore, directed to ascertain the prevalence and factors influencing satisfaction and to identify areas of dissatisfaction among caregivers at the Paediatric Specialist Clinic of Tunku Azizah Hospital. This newly established hospital is a tertiary facility and a national referral centre for the paediatric and women population.

## MATERIALS AND METHODS

### Subjects

This cross-sectional study was conducted at the Tunku Azizah Hospital, Kuala Lumpur, Malaysia. Subjects were caregivers to children seen with an appointment at the clinic. Exclusion criteria were (1) foreign nationals, (2) refusal to participate, (3) cognitively unsound, (4) caregivers who could not read, and (5) patient visiting for the first time. Only participants who met the eligibility criteria and agreed to participate in the study were enrolled. A total of 600 questionnaires distributed to the clinic, and we received 502 responses, giving a rate of 83.7%. Of these 502 responses, 43 were unusable and were excluded from this study, and the remaining 459 (91.4%) questionnaires were analysed. Some 2,238 patients were registered for an appointment at the clinic during this data collection period.

### Data Collection

This study was conducted at the hospital’s Paediatric Specialist Clinic by convenience sampling. Self-administered, structured questionnaires were distributed to consenting participants. Subjects who agreed to participate were given questionnaires after seeing the doctor and while waiting for the date of their next consultation. Participants were recruited for seven working days from 3rd to 12th September 2019 upon receiving approval from MREC. The principal researcher and two nurses were responsible for this data collection.

This was part of a hospital-level survey assessing satisfaction among caregivers attending the clinic using the SERVQUAL instrument. SERVQUAL was initially developed for use in the marketing industry (Parasuraman, Zeithaml and Berry, 1988). The SERVQUAL model is also known as a gap analysis model and is the most excellent tool for evaluating the quality of services (Brooks, Lings and Botschen, 1999). The analysis of gaps is based on the difference between service quality expectations and perception. It was modified, translated, and validated in line with the Malaysian healthcare setting (Roslan, 2002).

There are nine dimensions in this SERVQUAL tool, which includes the five original characteristics: tangibles, reliability, responsiveness, assurance, and empathy. Service outcomes, as well as three other dimensions, were included, which are the core values of MoH corporate culture: caring service, teamwork, and professionalism (John, Yatim and Mani, 2011). The current SERVQUAL tool that is used by MoH is phrased in dual language (Malay and English).

The first part of the survey, which addressed the demographics of the respondents, was modified to include demographics of paediatric patients visiting the clinics. Sociodemographic data included independent variables such as the caregivers’ age, gender, race, marital status, education, employment sector, and household income level. These followed by age and gender of the child (patient), relationship with the caregiver, sub specialty that is being visited, waiting time, and the main problem encountered at the clinic during their visit.

The second section included the SERVQUAL tool, which contains 16 statements related to the respondents’ expectations on quality of service and 18 statements concerning their perception (experience) with the quality of service delivered. A 5-point Likert Scale was used, ranging from “strongly disagree” (1) to “strongly agree” (5), with no verbal labels for scale points 2 through 4. Upon completing the questionnaire, participants were instructed to put it into an enclosed envelope. The sealed envelope is then passed to the nurse at the clinic counter.

### Statistical Analysis

The data were coded, entered into Microsoft Excel, and analysed using Statistical Package for the Social Sciences (SPSS) version 21 (IBM SPSS Inc.). Primary data on 459 responses were analysed to examine satisfaction with services provided at the clinic. Sociodemographic characteristics of caregivers and patients and patient’s clinic visits were analysed using descriptive analysis.

Each component from the satisfaction questionnaire was analysed using a chi-square test. To describe caregivers’ and patient demographic profiles, a descriptive model with frequencies and percentages were developed. The median score of expectations and perceptions of caregivers and the mean gap scores for 16 paired items were evaluated. The difference in the mean values of perception and expectation for each component determined the caregiver’s satisfaction. This methodology assesses service quality by measuring the discrepancy (gap) between caregivers’ perceptions and expectations (service quality = P–E). “P” reflects the perception of the caregivers, and “E” refers to expectations of service delivery before encountering the actual service (Berry, Shostack and Upah, 1983, pp.99– 107; Parasuraman, Zeithaml and Berry, 1985). If the difference is negative, then there is dissatisfaction. To evaluate the mean satisfaction gap for each dimension, the mean gaps from all statements pertaining to a dimension is summed and then divided by the number of statements making up that dimension.

The scores of four and five were considered to be satisfied, and the percentages were determined, while the other scores were considered to be dissatisfactory for the expectations and perception components. The Wilcoxon signed ranked test was used to make a comparison of distributions of expectations and perceptions. A logistic regression model was used to estimate the odds ratio (OR) and 95% confidence interval (CI) for overall satisfaction level. The mean was computed for all gap scores of 16 paired statements, and an average of zero and higher is considered satisfied. A p-value of less than 0.05 was considered to be statistically significant.

## RESULTS

Table 1 highlights the socio-demographic characteristics of respondents and patients. The total study population consisted of 144 males and 315 females (68.6%), with a significant proportion between 30 and 39 years of age (55.4%). Most respondents were from the Malay community (74.7%), and most were married (88.9%). About half of the respondents received a tertiary education (50.3%). Approximately one-third of the respondents were private-sector employees, and more than half of the respondents were from the lower category of household income. A vast majority of respondents were parents (94.1%), with more than half of the patients visiting were less than 60 months of age.

**Table 1.**
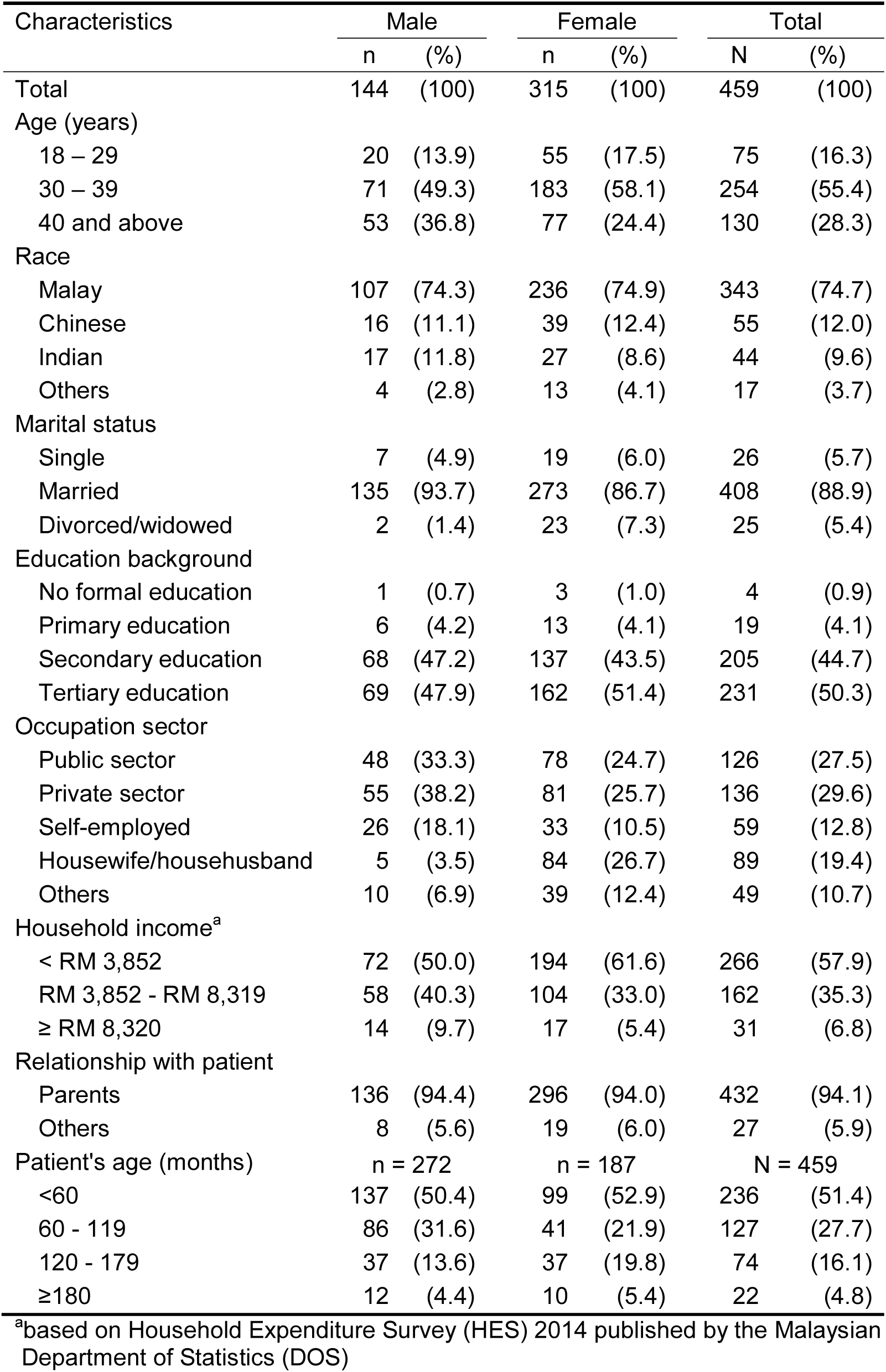
Socio-demographic characteristics of caregivers and patients (N=459)

| Characteristics | Male |  | Female |  | Total |  |
| --- | --- | --- | --- | --- | --- | --- |
|  | n | (%) | n | (%) | N | (%) |
| Total | 144 | (100) | 315 | (100) | 459 | (100) |
| Age (years) |  |  |  |  |  |  |
| 18 – 29 | 20 | (13.9) | 55 | (17.5) | 75 | (16.3) |
| 30 – 39 | 71 | (49.3) | 183 | (58.1) | 254 | (55.4) |
| 40 and above | 53 | (36.8) | 77 | (24.4) | 130 | (28.3) |
| Race |  |  |  |  |  |  |
| Malay | 107 | (74.3) | 236 | (74.9) | 343 | (74.7) |
| Chinese | 16 | (11.1) | 39 | (12.4) | 55 | (12.0) |
| Indian | 17 | (11.8) | 27 | (8.6) | 44 | (9.6) |
| Others | 4 | (2.8) | 13 | (4.1) | 17 | (3.7) |
| Marital status |  |  |  |  |  |  |
| Single | 7 | (4.9) | 19 | (6.0) | 26 | (5.7) |
| Married | 135 | (93.7) | 273 | (86.7) | 408 | (88.9) |
| Divorced/widowed | 2 | (1.4) | 23 | (7.3) | 25 | (5.4) |
| Education background |  |  |  |  |  |  |
| No formal education | 1 | (0.7) | 3 | (1.0) | 4 | (0.9) |
| Primary education | 6 | (4.2) | 13 | (4.1) | 19 | (4.1) |
| Secondary education | 68 | (47.2) | 137 | (43.5) | 205 | (44.7) |
| Tertiary education | 69 | (47.9) | 162 | (51.4) | 231 | (50.3) |
| Occupation sector |  |  |  |  |  |  |
| Public sector | 48 | (33.3) | 78 | (24.7) | 126 | (27.5) |
| Private sector | 55 | (38.2) | 81 | (25.7) | 136 | (29.6) |
| Self-employed | 26 | (18.1) | 33 | (10.5) | 59 | (12.8) |
| Housewife/househusband | 5 | (3.5) | 84 | (26.7) | 89 | (19.4) |
| Others | 10 | (6.9) | 39 | (12.4) | 49 | (10.7) |
| Household income<sup>a</sup> |  |  |  |  |  |  |
| < RM 3,852 | 72 | (50.0) | 194 | (61.6) | 266 | (57.9) |
| RM 3,852 - RM 8,319 | 58 | (40.3) | 104 | (33.0) | 162 | (35.3) |
| ≥ RM 8,320 | 14 | (9.7) | 17 | (5.4) | 31 | (6.8) |
| Relationship with patient |  |  |  |  |  |  |
| Parents | 136 | (94.4) | 296 | (94.0) | 432 | (94.1) |
| Others | 8 | (5.6) | 19 | (6.0) | 27 | (5.9) |
| Patient's age (months) | n = 272 |  | n = 187 |  | N = 459 |  |
| <60 | 137 | (50.4) | 99 | (52.9) | 236 | (51.4) |
| 60 - 119 | 86 | (31.6) | 41 | (21.9) | 127 | (27.7) |
| 120 - 179 | 37 | (13.6) | 37 | (19.8) | 74 | (16.1) |
| ≥180 | 12 | (4.4) | 10 | (5.4) | 22 | (4.8) |

<sup>a</sup>based on Household Expenditure Survey (HES) 2014 published by the Malaysian Department of Statistics (DOS)

Characteristics of patient’s clinic visits are presented in Table 2, which shows that the majority of them had four or more visits to the clinic by appointment (46.0%). Approximately two-thirds of the patients were seen by paediatric medical sub-specialties, and half of the total patients visited were seen by the physicians in less than 60 minutes.

**Table 2.** Characteristics of patient’s clinic visit (N=459)

| Characteristics | Male |  | Female |  | Total |  |
| --- | --- | --- | --- | --- | --- | --- |
|  | n | (%) | n | (%) | N | (%) |
| Total | 272 | (100) | 187 | (100) | 459 | (100) |
| Frequency of visit |  |  |  |  |  |  |
| 2 | 101 | (37.1) | 73 | (39.0) | 174 | (37.9) |
| 3 | 51 | (18.8) | 23 | (12.3) | 74 | (16.1) |
| ≥4 | 120 | (44.1) | 91 | (48.7) | 211 | (46.0) |
| Subspecialty visited |  |  |  |  |  |  |
| Paediatric medical | 173 | (63.6) | 131 | (70.1) | 304 | (66.2) |
| Paediatric surgical | 99 | (36.4) | 56 | (29.9) | 155 | (33.8) |
| Waiting time (minutes) |  |  |  |  |  |  |
| <60 | 138 | (50.7) | 92 | (49.2) | 230 | (50.1) |
| 60 - 119 | 61 | (22.4) | 38 | (20.3) | 99 | (21.6) |
| 120 - 179 | 50 | (18.4) | 44 | (23.5) | 94 | (20.5) |
| ≥180 | 23 | (8.5) | 13 | (7.0) | 36 | (7.8) |

Table 3 shows a comparison of expectation, perception, and satisfaction for each statement. This analysis shows that the respondents had a very high expectation for “staff politeness” (Q8), which was followed by “staff competency” (Q7) and “cleanliness of public toilets” (Q15). The least expectation was given for the “visual appeal of facilities” (Q2). However, it is interesting to note that the perception score fared slightly better for this statement. In terms of perception, the caregivers had the best experience with “staff politeness” (Q8), “staff work discipline” (Q13), and “cleanliness of public toilets” (Q15). On the contrary, the perception score was lowest for “staff providing services at promised time” (Q3) and “appropriate waiting time” (Q16). The highest satisfaction gap was observed with the “appropriate waiting time” (Q16) followed by the “staff providing services at the promised time” (Q3), and the lowest satisfaction gap was for the statement “visually appropriate physical facilities” (Q2).

**Table 3.** Comparison of distribution for expectation, perception, and satisfaction for each statement

| Measurement statements<sup>a</sup> | Expectation |  |  | Perception |  |  | Satisfaction gap |  | Z-statistic<sup>c</sup> | p-value<sup>c</sup> |
| --- | --- | --- | --- | --- | --- | --- | --- | --- | --- | --- |
|  | Score 4-5<sup>b</sup><br>(%) | Median | IQR | Score 4-5<sup>b</sup><br>(%) | Median | IQR | Mean | 95% CI |  |  |
| Q1 | 92.2 | 5 | 1 | 88.9 | 5 | 1 | -0.21 | -0.27, -0.14 | -6.08 | < 0.001 |
| Q2 | 89.8 | 5 | 1 | 90.0 | 5 | 1 | -0.07 | -0.13, 0.00 | -1.98 | < 0.050 |
| Q3 | 92.6 | 5 | 1 | 78.2 | 4 | 1 | -0.39 | -0.48, -0.30 | -8.29 | < 0.001 |
| Q4 | 93.7 | 5 | 1 | 87.8 | 4 | 1 | -0.26 | -0.33, -0.19 | -6.94 | < 0.001 |
| Q5 | 93.0 | 5 | 1 | 81.3 | 4 | 1 | -0.36 | -0.44, -0.28 | -7.88 | < 0.001 |
| Q6 | 93.7 | 5 | 1 | 89.3 | 5 | 1 | -0.21 | -0.27, -0.14 | -5.84 | < 0.001 |
| Q7 | 95.0 | 5 | 1 | 89.1 | 5 | 1 | -0.26 | -0.33, -0.19 | -6.88 | < 0.001 |
| Q8 | 95.2 | 5 | 1 | 91.3 | 5 | 1 | -0.21 | -0.28, -0.15 | -6.36 | < 0.001 |
| Q9 | 93.2 | 5 | 1 | 88.0 | 4 | 1 | -0.27 | -0.34, -0.19 | -6.68 | < 0.001 |
| Q10 | 92.4 | 5 | 1 | 88.7 | 4 | 1 | -0.23 | -0.31, -0.16 | -6.27 | < 0.001 |
| Q11 | 94.3 | 5 | 1 | 89.8 | 5 | 1 | -0.24 | -0.31, -0.17 | -6.53 | < 0.001 |
| Q12 | 93.7 | 5 | 1 | 89.3 | 5 | 1 | -0.22 | -0.29, -0.15 | -6.12 | < 0.001 |
| Q13 | 94.8 | 5 | 1 | 90.4 | 5 | 1 | -0.24 | -0.30, -0.17 | -6.83 | < 0.001 |
| Q14 | 94.3 | 5 | 1 | 88.7 | 4 | 1 | -0.27 | -0.34, -0.20 | -7.26 | < 0.001 |
| Q15 | 95.0 | 5 | 1 | 90.4 | 5 | 1 | -0.24 | -0.31, -0.17 | -6.44 | < 0.001 |
| Q16 | 92.8 | 5 | 1 | 79.3 | 4 | 1 | -0.48 | -0.57, -0.39 | -9.35 | < 0.001 |

<sup>a</sup> Refer appendix section for measurement statements.<sup>b</sup> Caregivers scoring 4 and 5.<sup>c</sup> Wilcoxon signed rank test.
Abbreviations: IQR, interquartile range; CI, confidence interval.

Table 4 depicts a comparison of expectation, perception, and satisfaction for each dimension. The “outcome” dimension had the most expectation from the caregivers, then by the “assurance” dimension. The lowest expectation was scored the “caring service” dimension. The caregivers’ perception was highest for the “outcome” dimension, as well. The “reliability” dimension had the lowest perception score and the widest satisfaction gap. The “tangibles” dimension, on the other hand, had the smallest satisfaction gap.

**Table 4.** Comparison of distribution for expectation, perception, and satisfaction for each dimension

| SERVQUAL dimensions<sup>a</sup> | Expectation |  |  | Perception |  |  | Satisfaction gap |  | Z-statistic<sup>c</sup> | p-value<sup>c</sup> |
| --- | --- | --- | --- | --- | --- | --- | --- | --- | --- | --- |
|  | Score 4-5<sup>b</sup><br>(%) | Median | IQR | Score 4-5<sup>b</sup><br>(%) | Median | IQR | Mean | 95% CI |  |  |
| Tangibles | 90.4 | 5.00 | 0.67 | 84.7 | 4.33 | 1.00 | -0.17 | -0.22, -0.12 | -6.15 | < 0.001 |
| Reliability | 90.4 | 5.00 | 0.67 | 75.4 | 4.33 | 1.00 | -0.35 | -0.44, -0.30 | -9.69 | < 0.001 |
| Responsiveness | 92.6 | 5.00 | 1.00 | 81.7 | 4.50 | 1.00 | -0.28 | -0.35, -0.21 | -7.58 | < 0.001 |
| Assurance | 93.7 | 5.00 | 0.67 | 85.6 | 4.67 | 1.00 | -0.25 | -0.31, -0.19 | -8.07 | < 0.001 |
| Empathy | 91.7 | 5.00 | 1.00 | 86.1 | 4.50 | 1.00 | -0.25 | -0.32, -0.18 | -6.96 | < 0.001 |
| Outcome | 94.3 | 5.00 | 1.00 | 89.8 | 5.00 | 1.00 | -0.24 | -0.31, -0.17 | -6.53 | < 0.001 |
| Caring service | 89.1 | 5.00 | 0.71 | 77.8 | 4.43 | 1.00 | -0.27 | -0.33, -0.22 | -8.73 | < 0.001 |
| Teamwork | 92.6 | 5.00 | 1.00 | 86.3 | 4.50 | 1.00 | -0.24 | -0.30, -0.18 | -7.32 | < 0.001 |
| Professionalism | 90.6 | 5.00 | 0.75 | 77.6 | 4.50 | 1.00 | -0.27 | -0.33, -0.21 | -8.87 | < 0.001 |

<sup>a</sup> Refer appendix section for dimension statements.<sup>b</sup> Caregivers scoring 4 and 5.<sup>c</sup> Wilcoxon signed rank test.
Abbreviations: IQR, interquartile range; CI, confidence interval.

Crude and adjusted odds ratio (AOR) with 95% confidence interval (CI) of the factors associated with the overall satisfaction of caregivers with the quality of services provided are demonstrated in Table 5. The OR was adjusted to 11 factors listed in Table 5. Caregivers from the Indian community (AOR: 2.91, 95% CI: 1.37-6.18) and lower household income groups (AOR: 2.94; 95% CI: 1.87-4.64) were approximately three times more likely to express higher levels of satisfaction with paediatric clinic service quality. Besides, respondents from lower educational backgrounds (AOR: 3.58; 95% CI: 1.19–10.72) were almost four times as likely to be satisfied with the services they received. However, housewives/househusbands (AOR: 0.48; 95% CI: 0.25–0.90) seemed less likely to be satisfied with the services provided during their visit to the clinic.

**Table 5.**
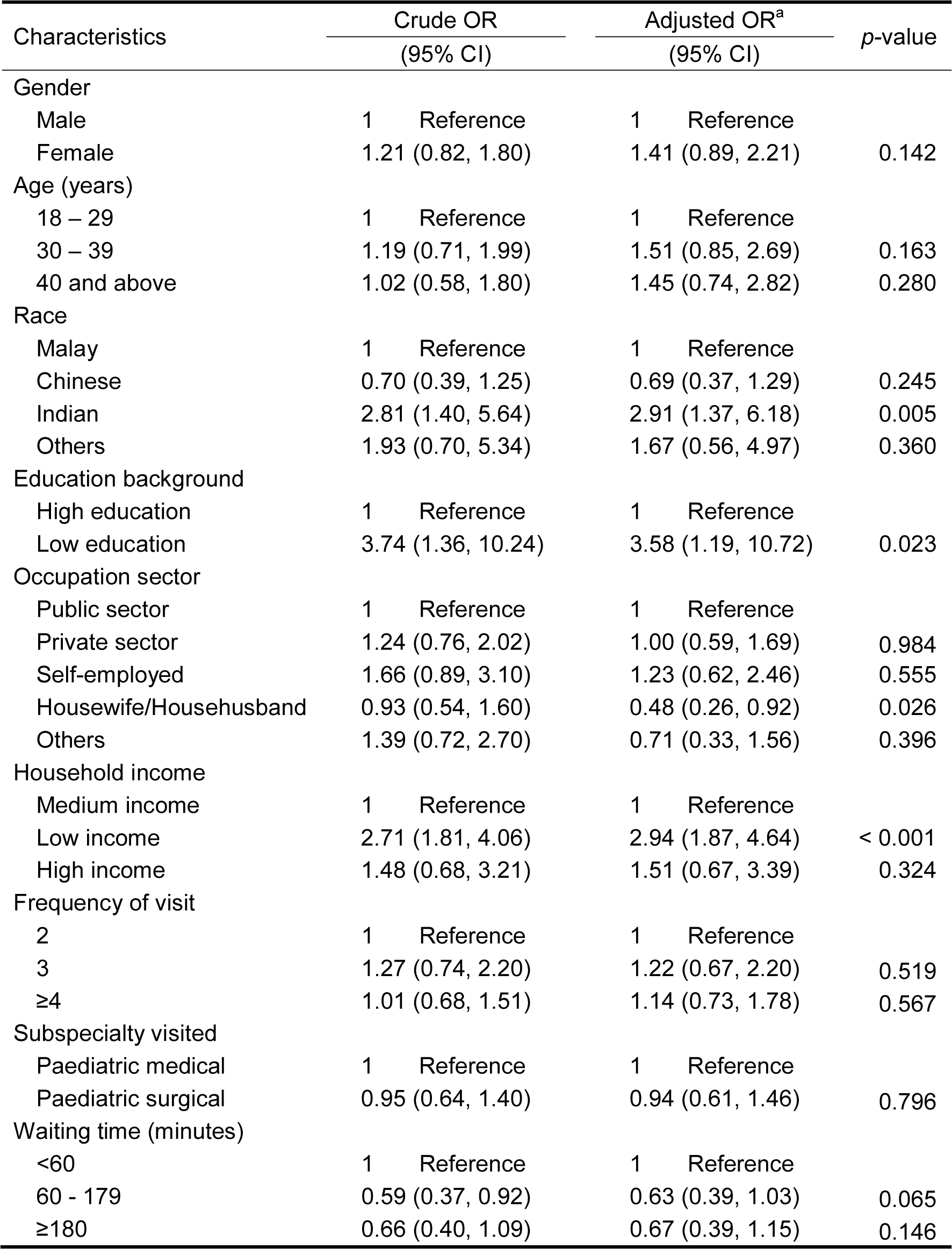
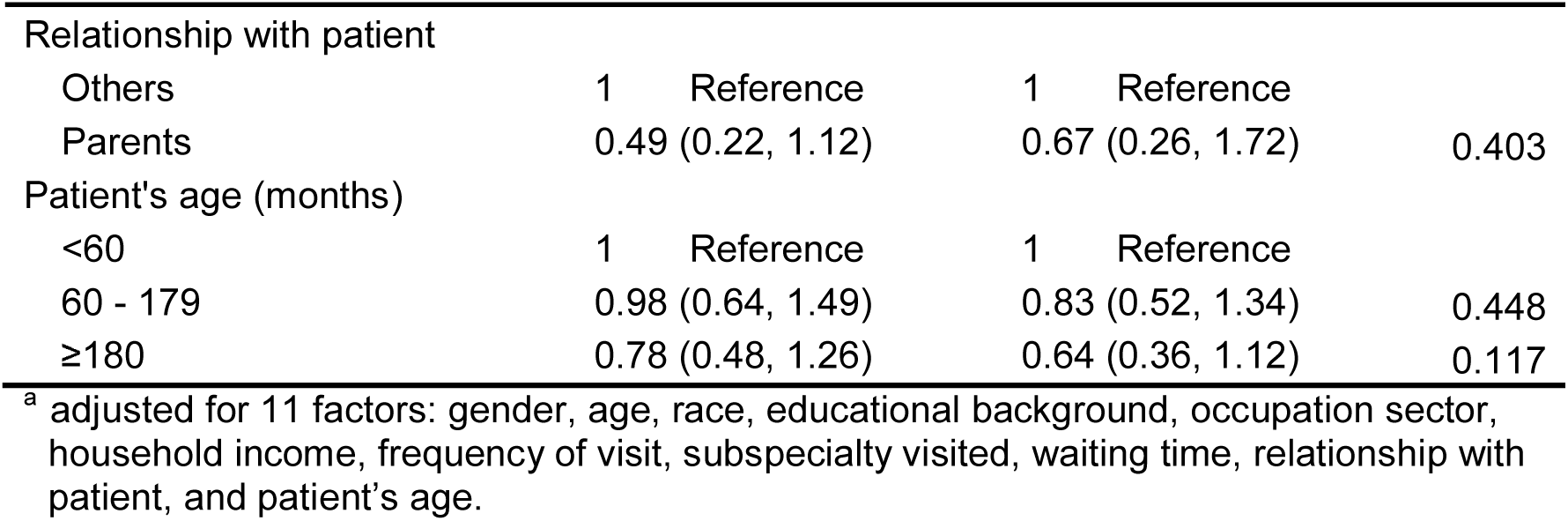
Odds ratio (OR) and 95% confidence interval (CI) of factors associated with the level of caregiver’s satisfaction (N=459)

Looking into the overall satisfaction of the patients with the quality of service encountered at the Paediatric Specialist Clinic, it can be derived that 50.5% (n=232) of caregivers demonstrated satisfaction with the quality of services, as opposed to 49.5% (n=227) of unsatisfied respondents.

## DISCUSSIONS

Tunku Azizah Hospital is the first Public-Private-Partnership (PPP) project in Malaysia under the MoH, utilizing the Private Finance Initiative (PFI) model. This facility was initially known as the Kuala Lumpur Women and Children Hospital but was renamed in January 2020 to commemorate the present Queen. The hospital started operations in phases from February 2019, and the Paediatric Specialist Clinic was the first to offer its services to the public. To our very best knowledge, this is the first paper that discusses the factors affecting overall caregivers’ satisfaction and identifies areas of dissatisfaction in a paediatric clinic run by MoH in Malaysia.

As can be seen from the results of the analysis, there was a negative satisfaction gap in all dimensions, suggesting that none surpassed the expectations of the caregivers. This result is also consistent with another research carried out in Singapore (Lim and Tang, 2000) using a similar instrument. Negative gaps are commonly predicted, as expectations for optimum service are rarely met.

Overall, 50.5% of caregivers were satisfied with the paediatric clinic and the quality of services provided during this study period. This result is in contrast with another study by Aniza et al (2009) which was conducted at the Paediatric Clinics of the University of Kebangsaan Malaysia Medical Center (UKMMC) that had a 90.5% satisfaction rate. Such a finding may be due to the higher expectations that caregivers had with a newly opened healthcare facility.

There was evidence that respondents with lower educational levels and household income, and ethnicity have better satisfaction with healthcare services at the clinic. Several authors have found that demographic characteristics, such as gender, age, and education, were strongly linked to respondent’s satisfaction. Although satisfaction levels were not significantly associated between age and gender in this study, their prevalence in other studies was significant, where males were found to be more satisfied than female respondents (Nguyen Thi et al., 2002). Another research indicated that gender did not have a significant impact on the satisfaction rate in their findings (Gajovic et al., 2012). Even if age does not appear to be associated with satisfaction levels in some research (Jaipaul and Rosenthal, 2003), one study found that the average satisfaction rate improved with the increase in age and that the satisfaction rate was the most feasible with those over 55 years (Afzal et al., 2014).

The current research found a statistically significant inverse association between the level of education and the satisfaction of caregivers, which is comparable to other studies indicating that respondents who were less educated were more satisfied than those with higher education (Hall and Dornan, 1990; Wallin et al., 2000; Minnick et al., 1997). This result could be due to higher standards set by educated group, as they believed they were more acquainted with the care they would obtain. Besides, those with higher education levels are pragmatic and able to see the services objectively, and they were dissatisfied when the level of services did not meet their expectation (Berry, Shostack and Upah, 1983, pp.99–107).

Similar to other research done, this study also shows that a lower income group has shown more satisfaction towards the services at the clinic (Ganasegeran et al., 2015). This group of caregivers consisted more than half of the total respondents and were more concerned about the costs associated with healthcare delivery. Thus, they were more satisfied with the services at this outpatient facility accessible at a low cost of RM 1.00. In this study, caregivers from the Indian ethnic minority were more satisfied, as opposed to the other ethnic groups. This is different from another article which pointed out that the minority ethnicity reported lower satisfaction and less positive experiences with health care services (Pinder, Ferguson and Møller, 2016).

One interesting observation from this study is that the housewives/househusbands had a relatively lower satisfaction level at the clinic, which is similar to another published article (Tateke, Woldie and Ololo, 2012). This could be due to the different commitments they have made, and they anticipate that the appointment will be completed in a short period of time.

In this research, all statements and dimensions revealed negative satisfaction scores indicating that none met the expectations of the caregivers. However, caregivers’ experiences from this study point out that the staff in the clinic had shown politeness, displaying good work discipline, and that the public toilets were clean. Caregivers were least satisfied with the waiting time and had concerns with services not being provided at the promised time. A study conducted in France also suggested dissatisfaction among patients with waiting times (Pitrou et al., 2009). MoH Malaysia had targeted 90 minutes of waiting time. Nevertheless, almost half of the patients (median waiting time) were seen by doctors in less than 60 minutes or at an average of 83 minutes for all cases.

The respondents also pointed out better than expected experience with the visual appeal of the healthcare infrastructure. As this facility is a PPP project, it has integrated certain non conventional elements into its architecture and design that reflect socio-cultural, economic, professional, and aesthetic priorities. This reflects and reinforces contemporary concepts of patient-hood and caring and projects the implementation of patient-centeredness.

Caregivers’ satisfaction with services can be assessed based on the following service attributes as highlighted by Parasuraman et al. (1988). The five original SERVQUAL dimensions are defined as reliability: the ability to perform the promised service dependably and accurately; responsiveness: willingness to help customers and provide prompt service; assurance: employees’ knowledge and courtesy and their ability to inspire trust and confidence; empathy: caring, individualized attention given to customers; and tangibles: the appearance of physical, facilities, personnel, and written materials. Additional four dimensions; service outcome, caring service, teamwork, and professionalism were included in the MoH version of SERVQUAL.

Caregivers had the highest expectation for service outcomes, and they also had the best experience with the outcome of their visits to the outpatient clinic, which indicates that they were pleased with consultations or treatments they got. However, the “reliability” dimension needs to be significantly enhanced, as this had the most substantial satisfaction gap. The care providers should focus on reducing the waiting time in the clinic and mobilizing resources to enhance customer satisfaction further. While the “tangibles” dimension had the lowest satisfaction gap over all other dimensions, it is equally important to clean, maintain, and gleam the building premises. Maintaining the building premises is essential to maintain the properties and protect the inhabitants of the building. Proper building maintenance ensures that the building and the environment remain a secure, clean, and safe to function.

There were some limitations to this study. Firstly, we carried out our study at a tertiary, national referral centre run by consultants, trained specialists, and post-graduate trainees, which differs from those in primary public clinics, which are mostly run by medical officers without postgraduate qualifications. Therefore, the results of our study cannot be generalized to reflect the performance of other clinics in this region. Since the questionnaires used were self-administered, illiterates were not recruited. Besides, convenience sampling, while unavoidable, is another drawback to this research due to the high probability of bias in sampling. Hence, the findings may not be generalized to the broader population. Also, not all aspects of the services, such as pharmacy and prescription drugs, have been evaluated in this study. These factors have been found to influence patient satisfaction significantly (Geitona et al., 2008; MartÍnez-López-de-Castro et al., 2018). This study was also carried out at a relatively new facility, which could have resulted in a positive satisfaction bias among some respondents.

We believe that future surveys with questionnaires should avoid using all positively expressed statements to assess service quality. It would mitigate the overall bias if there were a combination of positive and negative framed statements (Dunsch et al., 2018). Additionally, other aspects of services, such as registration, pharmacy, and prescription drugs, should be considered to gauge the complete experience of caregivers while visiting health facilities. Value-driven outcome tools that measure quality and include both nationally accepted and validated measures, as well as local physician and patient-defined outcomes measures, should also be considered (Lee et al., 2016).

## CONCLUSION

In conclusion, the lower income group, Indian ethnicity, and less educated respondents were more appreciative of the services provided by the Paediatric Specialist Clinic. We also did not find any statistical significance in satisfaction between patients of different age groups and gender. Regardless, we need to keep improvement measures in place to further boost the satisfaction among patrons, which will strengthen the standards of healthcare delivery. The hospital management will benefit from the incorporation of patient-centred care as a strategic investment goal, as well as the creation and execution of constructive, organized strategies that involve front-line clinicians in the process of improving caregiver’s satisfaction. Routine assessment of satisfaction should be done by using improvised questionnaires or other proven methods to identify unsatisfactory domains that need drastic improvements. These measures will ensure that the services provided are consistent with the MoH’s mission to provide quality and integrated people-centred health services to the masses.

## Data Availability

The data that support the findings of this study are available on request from the corresponding author.

## ACKNOWLEDGEMENT

We would like to express sincere appreciation and deep gratitude to Tunku Azizah Hospital, the Ministry of Health Malaysia, and Nagoya University for enabling this study to progress and for their involvement at every stage of this article. We would also like to thank the Director-General of Health Malaysia for his permission to publish this article.

## APPENDIX

Measurement statements and dimensions concerning satisfaction questionnaire used in the study.

| Statements |  |
| --- | --- |
| Q1 | Up-to-date equipment |
| Q2 | Visually appealing physical facilities |
| Q3 | Staff provides services at the promised time |
| Q4 | Staff render the services right, every time |
| Q5 | Patients can expect prompt service from the staff |
| Q6 | Staff always willing to help with sincere interest |
| Q7 | Staff do their job competently |
| Q8 | Staff are polite |
| Q9 | Staff always understand patients' needs |
| Q10 | Staff render patients personal attention |
| Q11 | Staff provide effective treatment |
| Q12 | Staff work as a team in rendering treatment |
| Q13 | Staff display good work discipline |
| Q14 | Staff are knowledgeable |
| Q15 | Public toilets are always clean |
| Q16 | Waiting time is appropriate |
| Q17 | Staff provide services in accordance with the ministry's client charter |
| Q18 | Overall level of satisfaction |
| Dimensions |  |
| <u>SERVQUAL dimensions</u> |  |
| Tangibles | Q1, Q2, Q15 |
| Reliability | Q3, Q4, Q16 |
| Responsiveness | Q5, Q6 |
| Assurance | Q7, Q8, Q14 |
| Empathy | Q9, Q10 |
| Outcome | Q11 |
| <u>Ministry of Health corporate culture core value dimensions</u> |  |
| Caring service | Q3, Q4, Q5, Q6, Q8, Q9, Q10 |
| Teamwork | Q7, Q12 |
| Professionalism | Q3, Q4, Q7, Q9, Q10, Q11, Q13, Q14 |

